# Establishing Normative Measurements and Ratios for Rotator Cuff Muscles in a Military Population Using Diagnostic Ultrasound

**DOI:** 10.1101/2025.08.12.25333295

**Authors:** Jordan E. Powell, Megan R. Loftsgaarden, Marin S. Smith, Alexis Southwell, Cristina Abboud Chalhoub, Kathryn Roberts, Riley R. Boeth, Sean R. Wise, Xiaoning Yuan

## Abstract

**Introduction:** Rotator cuff (RTC) muscle size is predictive of shoulder function and potential pathology following injuries. Ultrasound (US) can differentiate normative from pathologic RTC musculature, and inform the diagnosis of shoulder pathology and treatment selection for more efficient, patient-centered care. This study’s objective was to establish normative sonographic measurements and ratios for RTC musculature within a military population.

**Methods:** In this cross-sectional observational study at three military treatment facilities, military beneficiaries underwent a standardized US scanning protocol to measure the upper trapezius (UT), supraspinatus (SS), infraspinatus (IS), and teres minor (TM) muscle dimensions. Ratios of these dimensions were calculated, compared to adjacent muscles.

**Results:** 128 participants (52 female; 88.2% active duty, mean age: 32.9 years; 231 shoulders) were enrolled, excluding shoulders with a history of significant injury or surgery. Mean and standard deviation (SD) for UT and SS thickness (mm) were 8.8 ± 2.2 and 22.2 ± 4.1, respectively. Mean and SD for SS, IS, and TM cross-sectional area (CSA; cm^2^) were 6.48 ± 1.82, 8.65 ± 2.79, and 4.19 ± 1.45, respectively. Ratios (mean ± SD) for UT:SS thickness, SS:IS CSA, and IS:TM CSA were as follows: 0.41 ± 0.13, 0.77 ± 0.19, 2.17 ± 0.73. Subgroup analyses demonstrated significantly larger dominant-sided RTC muscle dimensions for biological males than females (*p* < 0.01), but corresponding ratios were not significantly different.

**Conclusion:** This is the largest US study to date to examine RTC musculature, establishing normative sonographic measurements and ratios in a military population. Future research should determine if RTC muscle ratios are more predictive than absolute dimensions of shoulder pathology in symptomatic personnel, to guide clinical decision-making and optimize warfighter readiness.

**Key Messages:**

- US offers a cost-effective, efficient, and validated alternative to MRI for assessing RTC muscle size, which can be predictive of shoulder function and pathology.
- This study established normative sonographic measurements and ratios for RTC muscle dimensions in an active, healthy, military population, providing a robust reference that accounts for demographic differences, such as biological sex, which can affect absolute muscle size.
- By introducing muscle dimension ratios as a diagnostic tool, this study enhances the potential for personalized assessment of shoulder health, which can enable more timely, accurate diagnosis and patient-centered treatment plans, ultimately optimizing warfighter readiness.
- Further research is needed to determine the predictive value of RTC muscle ratios for specific shoulder pathologies (e.g., RTC tendon tear, denervation), compared to absolute RTC muscle dimensions alone.

## Introduction

Active duty Service Members (ADSMs) undergo intense physical demands in both training and operational environments, resulting in musculoskeletal injuries that represent the top threat to medical readiness. The largest proportion of upper extremity injuries within the U.S. military occurs in the shoulder, affecting 8.1% of ADSMs [1]. Considering the significant role that the RTC plays in shoulder stabilization, functional activity, and the physical fitness requirements for ADSMs, having the means to efficiently evaluate RTC integrity is critical.

Advanced imaging techniques can quantify RTC muscle size, which is predictive of both function and potential pathology [2]. The maximum cross-sectional area (CSA) of each RTC muscle is closely related to the muscle’s isokinetic strength [3], while diminished CSA of these muscles correlates with RTC pathology [4] and is a harbinger of poor surgical outcomes [5–7]. Following significant RTC tendon or nerve injury, reduced muscle size and fatty degeneration co-occur, and are predictive of deficits in strength and function, including loss of abduction, external rotation, and internal rotation [8,9]. The CSA of RTC muscles has been assessed by magnetic resonance imaging (MRI) for both asymptomatic and symptomatic RTCs [2,4,10–13]. However, MRI is time-consuming, expensive, and patients frequently experience lengthy wait times for access to studies, prompting the search for more cost-effective, efficient imaging alternatives that can be utilized as both diagnostic and screening tools.

Musculoskeletal ultrasound (MSK US) shows promise as a diagnostic adjunct to facilitate timely diagnosis and targeted treatments [14–16]. As current literature is inconsistent regarding the value of objective physical examination maneuvers and special tests, MSK US can serve as a valuable aid during evaluations [14,17], and be applied at the point-of-care in a variety of settings to assess the RTC musculature and diagnose shoulder pathology quickly and cost-effectively. While shoulder dysfunction has been linked with strength and motor control deficits of the RTC and periscapular musculature, literature suggests that current strength testing and movement analyses may have limitations in detecting injury risk within the military population, given their unique occupational demands [18,19]. As an adjunct, MSK US offers the potential to provide diagnostic information pertaining to musculotendinous integrity and functional capacity. Compared to MRI, US is less expensive and can be performed efficiently at bedside, in the clinic, and in military and pre-hospital settings [20], given provider proficiency in MSK US and access to an US system. Studies have demonstrated that US measurements of RTC muscle dimensions, such as CSA and thickness, have good correlation with MRI measurements [11,21]. Additionally, US has been used to examine the quality of RTC muscles, such as visualizing atrophy, fatty infiltration, or fibrosis, which also correlates well with MRI findings [12,22].

However, measuring absolute values for RTC muscle dimensions may have limited clinical utility. Although prior research showed an association between diminished supraspinatus (SS) muscle CSA and the presence of RTC tendon pathology, women and individuals with lower fat-free mass may have lower RTC muscle CSAs, when compared to men and individuals with higher fat-free mass [13]. Thus, on an individual level, absolute muscle dimensions may not be as reliable of a predictor of potential pathology across certain demographic groups. Using ratios may adjust for these differences and serve as promising, alternative diagnostic tools at the point-of-care [14]. The only ratio described in advanced imaging literature of the RTC is the occupation ratio, which compares the transverse SS CSA to the CSA of the supraspinatus fossa. Clinically, a reduced occupation ratio is a predictor of SS tendon tears and poor surgical outcomes [6,23]. US-based ratios of muscle CSA have also been validated in other parts of the body, such as the fibularis longus and fibularis brevis at the ankle. Calvo Lobo et al. compared the two muscle CSAs using a ratio and found lower CSA ratios for sprained ankles compared to healthy ankles [24].

Besides the occupation ratio, no other established ratios exist for the RTC muscles. Comparing a RTC muscle to an adjacent muscle as a ratio may have more clinical utility for individuals from different demographic groups, rather than comparing the absolute dimensions of their muscles alone. Therefore, the aims of this study are to: 1) establish normative reference values of sonographic measurements for the absolute dimensions (thickness, CSA) of RTC muscles (SS, IS, TM), 2) obtain normative ratios for RTC muscles relative to adjacent muscles, to include the upper trapezius (UT):SS thickness ratio, SS:IS CSA ratio, and the IS:TM CSA ratio, and 3) determine if age, arm dominance, or biological sex influence these measurements and ratios within an active, healthy, military population.

## Methods

### Setting

This was a cross-sectional observational study enrolling participants from April 1, 2021, to May 15, 2023, at three military treatment facilities (MTFs). Research approval was obtained through the primary MTF’s Institutional Review Board.

### Participants

Inclusion criteria for enrollment required participants to be between the ages of 18 to 50 (inclusive) and enrolled in the Defense Enrollment Eligibility Reporting System. Participants were seen within the Physical Medicine & Rehabilitation or Sports Medicine clinics at MTFs. After obtaining informed consent, participants were screened via patient and provider report for potential exclusion criteria: history of muscular dystrophy, myopathy, or motor neuron disease. Each participant’s eligible shoulders were then screened individually, excluding shoulders with a history of surgery, significant injury, nerve disorder (e.g., cervical radiculopathy, brachial plexopathy, suprascapular nerve injury), or currently receiving treatment for pathology. Prior to data collection, diagnostic US was performed on the eligible shoulders’ RTC tendons to evaluate for major, asymptomatic, structural pathology. Shoulders with a > 50% thickness tear of any RTC tendon were excluded. Given that partial-thickness tears are present among 36% of asymptomatic individuals, shoulders with ≤ 50% thickness tears were included [25].

### Data Collection

The following demographic information was collected by participant report: age, biological sex, race/ethnicity, history of shoulder injury and/or symptoms, arm dominance, military affiliation, employment status, and daily activities. Ultrasound data collection was performed exclusively by four authors (JP, ML, SW, XY), who are physicians with a combined 23 years of experience practicing MSK US. Three (JP, SW, XY) have Registered in Musculoskeletal Sonography (RMSK) certification. Images obtained by the non-RMSK-certified physician (ML) were reviewed for accuracy by a co-author who was RMSK-certified (JP). All examiners completed two training sessions to review scanning and measurement techniques. All US exams were performed with a GE Logiq e Nextgen using an 8-12 MHz linear transducer or a GE Logiq S8 using a 6-15 MHz linear transducer (Boston, MA). During the exam, participants were seated with their forearm in supination, resting on their ipsilateral thigh (**FIGURE 1A**). US measurements were performed immediately after examination or during post-hoc image review.

**FIGURE 1.**
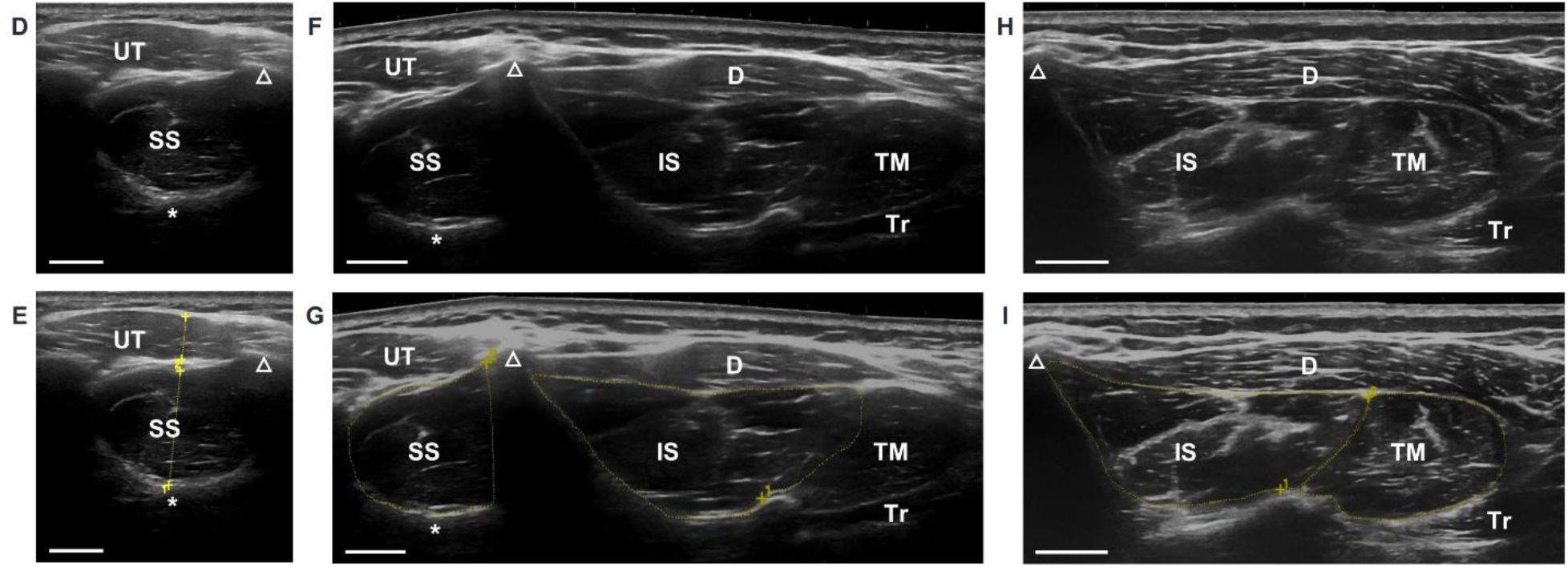
Participant and ultrasound (US) transducer positioning, with corresponding sample US images and measurements. Please contact the corresponding author to request access to the participant images (**A-C**). The participant is seated with their forearm in supination, resting on their ipsilateral thigh. The scapular spine is marked on the participant (**A**) and indicated by a dashed white line (**B**). Upper trapezius (UT) and supraspinatus (SS) images (**D**) were obtained in transverse axis with the transducer (**A**) at the level of the suprascapular notch (asterisk), as indicated by the green line (**B**). Corresponding UT and SS thickness measurements are presented as dashed yellow lines (**E**). Extended field-of-view (EFOV) SS and infraspinatus (IS) images (**F**) were obtained in transverse axis with the transducer following the trajectory delineated by the green line and red arrow (**B**), from the supraspinatus fossa at the level of the suprascapular notch to the medial border of the scapular attachment of the long head of the triceps (Tr). Corresponding SS and IS cross-sectional area (CSA) measurements are presented as dashed yellow lines (**G**). EFOV IS and teres minor (TM) images (**H**) were obtained in transverse axis with the transducer (**C**) following the trajectory delineated by the red arrow (**B**) from the suprascapular notch to the medial border of the Tr scapular attachment. After acquiring images, gain was increased as needed by sonographers to facilitate measurements. Corresponding IS and TM CSA measurements are presented as dashed yellow lines (**I**). Abbreviations: *D*, deltoid; Δ, scapular spine; *UT*, upper trapezius; *SS*, supraspinatus; *CSA*, cross-sectional area; *IS*, infraspinatus; *TM*, teres minor. Scale bar = 1 cm.

#### UT Thickness, SS Thickness, UT:SS Thickness Ratio

The US transducer was positioned to visualize the supraspinatus fossa, with the SS muscle in transverse axis (**FIGURE 1A, D**). In alignment with prior US studies of the SS muscle, the suprascapular notch was used as a landmark to ensure consistency in technique across participants [21,26]. SS thickness was measured by drawing a straight line between the suprascapular notch and perpendicular to the inner fascial border of the SS (**FIGURE 1E**). UT thickness was measured as a straight line between the superficial and deep inner fascial borders of the UT (**FIGURE 1E**). For each included shoulder, three unique images of the supraspinatus fossa were saved, with the transducer removed and repositioned to acquire each image. UT and SS thickness were measured per image using the US system. UT:SS thickness ratio was calculated per image during post-hoc data analysis. All values were reported as an average of the three unique images per shoulder.

#### SS CSA, IS CSA, SS:IS CSA Ratio

To capture extended field-of-view (EFOV) images of both the SS and IS for CSA measurement, the suprascapular notch was utilized again as a sonographic landmark to ensure consistency in technique across participants. The supraspinatus fossa at the level of the suprascapular notch was used as the superomedial boundary, while the medial aspect of the long head of the triceps’ scapular attachment served as the inferolateral boundary. An EFOV image was obtained, visualizing the SS and IS in the transverse axis (**FIGURE 1F**), by translating the transducer in a straight line between these two boundaries, while maintaining adequate visualization and image focus (**FIGURE 1B**). SS and IS CSAs were measured per image by tracing the borders of the respective muscles (**FIGURE 1G**) using the US system. For each included shoulder, three unique EFOV images were saved, with the transducer removed and repositioned to acquire each image. SS:IS CSA ratio was calculated per image during post-hoc data analysis. All values were reported as an average of the three unique images per shoulder.

#### IS CSA, TM CSA, IS:TM CSA Ratio

Similar to the approach for SS and IS, an EFOV image was obtained, visualizing the IS and TM in the transverse axis (**FIGURE 1C, H**). The suprascapular notch was again used as the superomedial boundary, while the medial aspect of the long head of the triceps’ scapular attachment served as the inferolateral boundary. The EFOV image was obtained by translating the transducer in a straight line between these two boundaries, while maintaining adequate visualization and image focus (**FIGURE 1B**). IS and TM CSAs were measured per image by tracing the borders of the respective muscles (**FIGURE 1I**) using the US system. For each included shoulder, three unique EFOV images were saved, with the transducer removed and repositioned to acquire each image. IS:TM CSA ratio was calculated per image during post-hoc data analysis. All values were reported as an average of the three unique images per shoulder, with the exception of IS CSA, which was reported as the average of six unique images captured per shoulder during EFOV imaging of the SS and IS, and the IS and TM.

### Statistical Methods

Statistical analyses were performed in the R programming language (Vienna, AT) and included measures of distribution to produce normative ranges, Z-scores to compare subgroups and evaluate specific subgroup differences, and Pearson’s correlations to evaluate the strength of relationships between measures. An α of 0.01 was chosen to reduce the risk of false discovery given the number of statistical tests performed. Each eligible shoulder’s absolute dimensions (UT thickness, SS thickness, SS CSA, IS CSA, TM CSA) and ratios (UT:SS thickness, SS:IS CSA, IS:TM CSA) were reported as the average of three unique images per shoulder, with the exception of IS CSA, which was reported as the average of six unique images captured per shoulder during EFOV imaging of the SS and IS, and the IS and TM. Absolute dimensions and ratios were then evaluated using means, standard deviations (SD), and 95% confidence intervals (CIs) across all shoulders included in the study. Subgroup analysis (arm dominance, biological sex) was performed for RTC measurements and ratios using two-tailed Z-scores. To avoid inappropriate analysis of paired data, only the measurements for dominant-sided shoulders were used when comparing biological males to females. Normality testing of data was performed using boxplots and/or histograms, prior to calculation of Pearson’s correlation coefficients to examine the relationship of numeric age with dominant-sided RTC measurements.

## Results

### Study Population

A total of 128 participants (76 male, 52 female, 88.2% active duty) were included. The mean age was 32.9 ± 7.5 years. Additional available demographic characteristics are listed in **TABLE 1**. 23 individual shoulders were excluded for the following reasons: history of shoulder surgery or significant shoulder injury, nerve injury involving the shoulder. Following their US exam, two asymptomatic shoulders were excluded from analysis, as their TM demonstrated sonographic evidence of atrophy and fibrosis, which may suggest prior nerve injury, although idiopathic TM atrophy has been previously described in literature [27]. Furthermore, 14 shoulders had their EFOV-based measurements (SS CSA, IS CSA, TM CSA, SS:IS CSA ratio, IS:TM CSA ratio) excluded after review of their images, because the medial aspect of the long head of the triceps’ scapular attachment was not visualized. Ultimately, a total of 231 shoulders were included in the study (**FIGURE 2**), including 217 shoulders with all sonographic measurements and ratios available for analysis.

**FIGURE 2.**
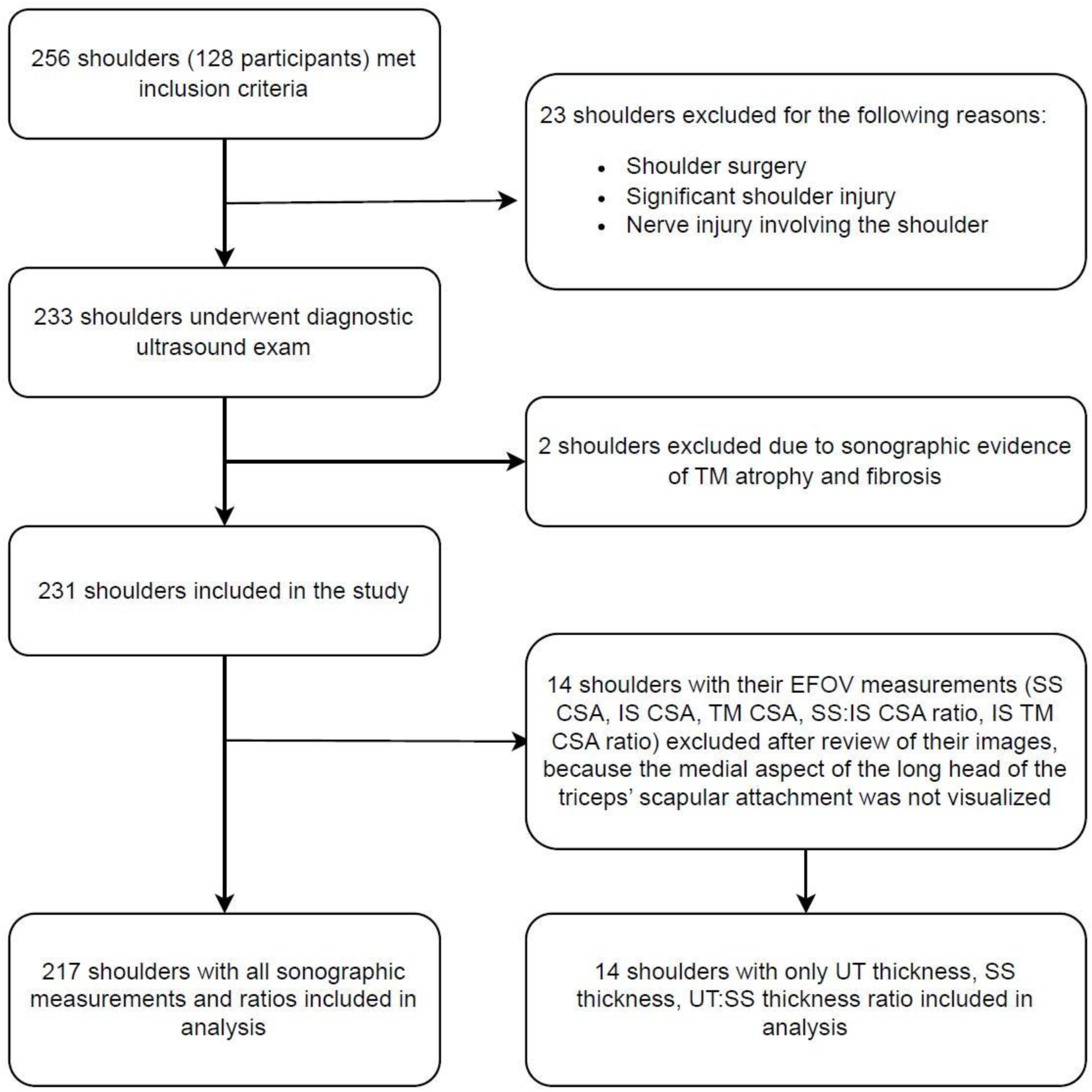
Study flow diagram of eligible participants and shoulders included in data analysis.

**TABLE 1.**
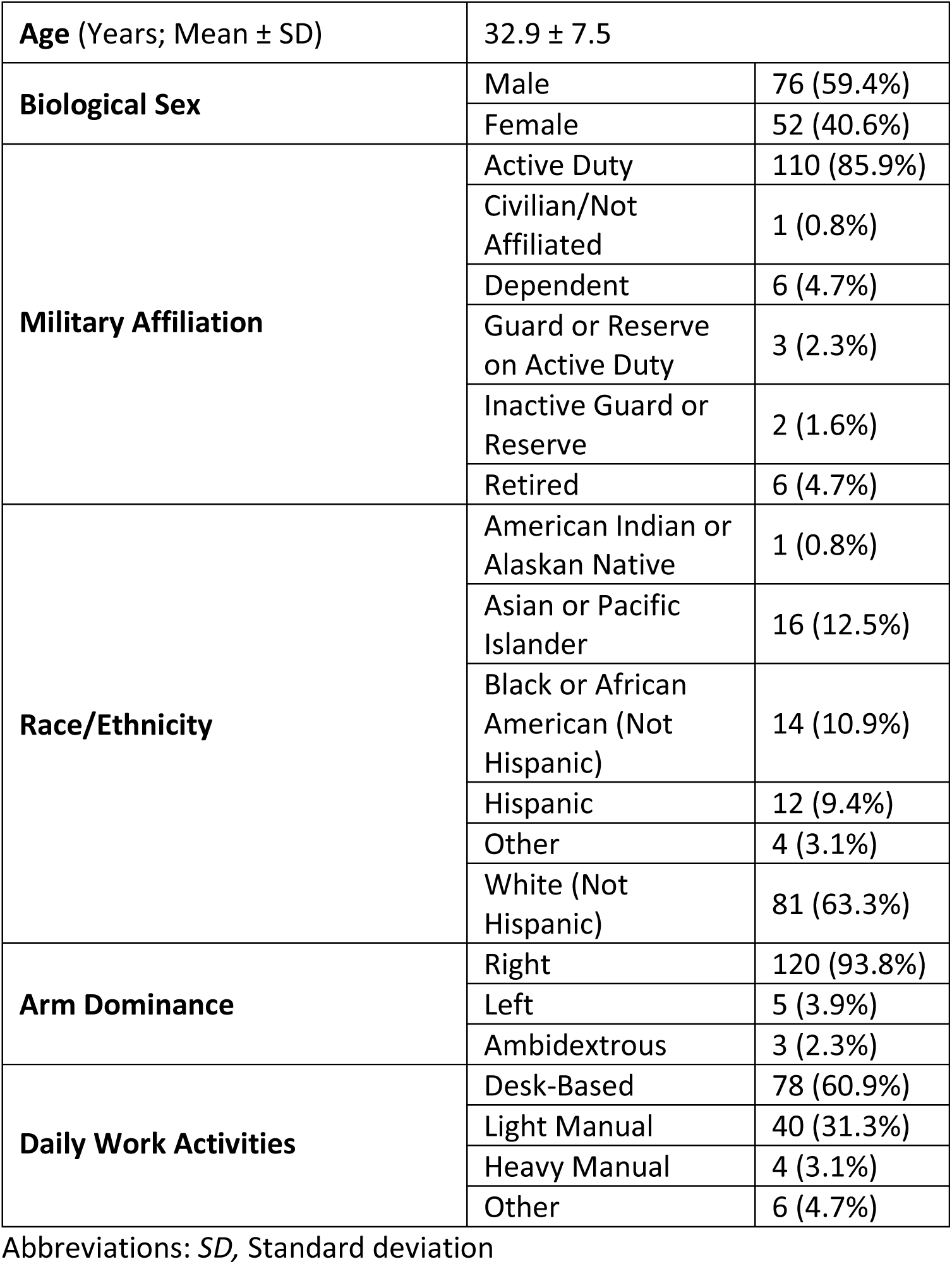
Demographic characteristics of study participants.

|  |  |  |
| --- | --- | --- |
| **Age (Years; Mean ± SD)** | 32.9 ± 7.5 | |
| **Biological Sex** | Male | 76 (59.4%) |
|  | Female | 52 (40.6%) |
| **Military Affiliation** | Active Duty | 110 (85.9%) |
|  | Civilian/Not Affiliated | 1 (0.8%) |
|  | Dependent | 6 (4.7%) |
|  | Guard or Reserve on Active Duty | 3 (2.3%) |
|  | Inactive Guard or Reserve | 2 (1.6%) |
|  | Retired | 6 (4.7%) |
| **Race/Ethnicity** | American Indian or Alaskan Native | 1 (0.8%) |
|  | Asian or Pacific Islander | 16 (12.5%) |
|  | Black or African American (Not Hispanic) | 14 (10.9%) |
|  | Hispanic | 12 (9.4%) |
|  | Other | 4 (3.1%) |
|  | White (Not Hispanic) | 81 (63.3%) |
| **Arm Dominance** | Right | 120 (93.8%) |
|  | Left | 5 (3.9%) |
|  | Ambidextrous | 3 (2.3%) |
| **Daily Work Activities** | Desk-Based | 78 (60.9%) |
|  | Light Manual | 40 (31.3%) |
|  | Heavy Manual | 4 (3.1%) |
|  | Other | 6 (4.7%) |

Abbreviations: *SD*, Standard deviation

### Quantitative Measurements

The mean, SD, 95% CIs, *p*-values, and number of included shoulders for each measurement and ratio is listed in **TABLE 2** across all participants and by subgroups (arm dominance, biological sex). Neither the mean RTC muscle dimensions nor ratios were significantly different between dominant and non-dominant arms. While biological females’ dominant-sided RTC muscles demonstrated significantly smaller dimensions (*p* < 0.01 for all thickness and CSA measurements), their corresponding ratios were not significantly different from their male counterparts. There was no significant correlation between age and dominant-sided RTC muscle dimensions or ratios, except for a weak positive correlation for the IS:TM CSA ratio with age [Pearson correlation coefficient: 0.25; *p* = 0.008; 95% CI (0.07, 0.42)].

**TABLE 2.** Normative sonographic measurements and ratios of rotator cuff and adjacent muscles: means, standard deviations (SDs), 95% confidence intervals (95% CIs), *p*-values, and number of eligible shoulders (N) per measurement and ratio for all participants and by subgroup (arm dominance, biological sex).

| Measurement / Ratio |  | All Shoulders | Arm Dominance |  | Biological Sex<sup>†</sup> |  |
| --- | --- | --- | --- | --- | --- | --- |
|  |  |  | Dominant | Non-Dominant | Male | Female |
| UT Thickness (mm) | Mean | 8.8 | 8.9 | 8.8 | 9.6* | 7.9* |
|  | SD | 2.2 | 2.1 | 2.2 | 1.9 | 2.0 |
|  | 95% CI | 8.5, 9.1 | 8.5, 9.3 | 8.4, 9.2 | 9.1, 10.1 | 7.3, 8.5 |
|  | *p*-value |  | 0.78 |  | 2.1×10<sup>-5</sup> |  |
|  | N | 231 | 113 | 113 | 65 | 48 |
| SS Thickness (mm) | Mean | 22.2 | 22.1 | 22.2 | 23.9* | 19.6* |
|  | SD | 4.1 | 4.1 | 4.2 | 3.9 | 3.0 |
|  | 95% CI | 21.7, 22.7 | 21.3, 22.9 | 21.4, 23.0 | 22.9, 24.9 | 18.7, 20.5 |
|  | *p*-value |  | 0.83 |  | 2.7×10<sup>-9</sup> |  |
|  | N | 231 | 113 | 113 | 65 | 48 |
| SS CSA (cm<sup>2</sup>) | Mean | 6.48 | 6.38 | 6.51 | 7.55* | 4.78* |
|  | SD | 1.82 | 1.78 | 1.87 | 1.10 | 1.17 |
|  | 95% CI | 6.23, 6.73 | 6.04, 6.72 | 6.14, 6.88 | 7.27, 7.83 | 4.43, 5.13 |
|  | *p*-value |  | 0.59 |  | 2.0×10<sup>-16</sup> |  |
|  | N | 217 | 109 | 103 | 63 | 46 |
| IS CSA (cm<sup>2</sup>) | Mean | 8.65 | 8.71 | 8.56 | 10.26* | 6.58* |
|  | SD | 2.79 | 2.86 | 2.78 | 2.47 | 1.79 |
|  | 95% CI | 8.27, 9.03 | 8.16, 9.26 | 8.01, 9.11 | 9.64, 10.88 | 6.05, 7.11 |
|  | *p*-value |  | 0.81 |  | 4.1×10<sup>-13</sup> |  |
|  | N | 217 | 109 | 103 | 63 | 46 |
| TM CSA (cm<sup>2</sup>) | Mean | 4.19 | 4.22 | 4.11 | 4.93* | 3.25* |
|  | SD | 1.45 | 1.47 | 1.45 | 1.39 | 0.93 |
|  | 95% CI | 3.99, 4.39 | 3.94, 4.5 | 3.82, 4.4 | 4.58, 5.28 | 2.98, 3.52 |
|  | *p*-value |  | 0.51 |  | 1.3×10<sup>-11</sup> |  |
|  | N | 217 | 109 | 103 | 63 | 46 |
| UT:SS Thickness Ratio | Mean | 0.41 | 0.41 | 0.41 | 0.41 | 0.41 |
|  | SD | 0.13 | 0.12 | 0.13 | 0.12 | 0.11 |
|  | 95% CI | 0.39, 0.43 | 0.39, 0.43 | 0.39, 0.43 | 0.38, 0.44 | 0.38, 0.44 |
|  | *p*-value |  | 0.69 |  | 0.89 |  |
|  | N | 231 | 113 | 113 | 65 | 48 |
| SS:IS CSA Ratio | Mean | 0.77 | 0.76 | 0.78 | 0.77 | 0.74 |
|  | SD | 0.19 | 0.19 | 0.18 | 0.21 | 0.16 |
|  | 95% CI | 0.74, 0.80 | 0.61, 0.91 | 0.65, 0.91 | 0.72, 0.82 | 0.69, 0.79 |
|  | *p*-value |  | 0.56 |  | 0.65 |  |
|  | N | 217 | 109 | 103 | 63 | 46 |
| IS:TM CSA Ratio | Mean | 2.17 | 2.19 | 2.17 | 2.27 | 2.07 |
|  | SD | 0.80 | 0.80 | 0.67 | 0.96 | 0.47 |
|  | 95% CI | 2.15, 2.23 | 2.15, 2.23 | 2.13, 2.21 | 2.03, 2.51 | 1.93, 2.21 |
|  | *p*-value |  | 0.80 |  | 0.68 |  |
|  | N | 109 | 109 | 103 | 63 | 46 |

Abbreviations: *UT*, upper trapezius; *SS*, supraspinatus; *CSA*, cross-sectional area; *IS*, infraspinatus; *TM*, teres minor
<sup>†</sup>Dominant-sided shoulders only
\* $p < 0.01$ between subgroups

## Conclusions

This is the largest US study to date that examines the RTC musculature, and the first to report the ratios of RTC muscle dimensions relative to adjacent musculature. In concordance with previous studies, biological males had significantly larger RTC dimensions compared to females. The sonographic CSAs of the individual muscles were overall in concordance with prior MR and US-based studies, despite differences in examination technique, imaging modality, and patient population [10,13,21,28].

Variance between individuals, such as biological males and females, presents a diagnostic challenge to determine if a specific RTC muscle dimension constitutes potential pathology. Our results support the potential clinical utility of a ratio comparing the dimensions of adjacent muscles to adjust for differences in RTC muscles between men and women. While absolute RTC dimensions were significantly different between men and women, their ratios were similar. Furthermore, we found no difference in normative measurements and ratios between dominant and non-dominant shoulders, which aligns with Karthikeyan et al.’s findings that suggest an asymptomatic, contralateral shoulder may serve as a control during the US examination of a symptomatic shoulder [29].

There was no correlation between dimensions or ratios and age, with one exception: the IS:TM ratio showed a weak positive correlation with age [Pearson correlation coefficient: 0.25; *p* = 0.008; 95% CI (0.07, 0.42)]. This may relate to the known increased incidence of TM atrophy with age [27].^7^

The examiners noted that in a minority of patients, it can be challenging to distinguish the borders of the IS and TM. The fascia demarcating these two muscles cannot be easily distinguished in a minority of individuals, although dynamic scanning from medial to lateral in the transverse axis can aid in visualizing the respective muscles’ borders, which become myotendinous and more distinct. Typically, a small fascial septum that separates these two muscles at their superficial aspect can be visualized [30], along with the hyperechoic bony ridge on the scapula that separates the deep aspects of these muscles. The fascia separating the IS and TM can then be traced between the superficial fascial septum and the ridge on the scapula.

There are several limitations to our study. First, the normative sonographic measurements and ratios were all captured in a two-dimensional plane at specified locations within each muscle and therefore are not representative of cumulative RTC muscle sizes, dimensions, or ratios. Additionally, it is important to note that all our study participants were military beneficiaries, including ADSMs, which may limit generalizability to the general population. Our data does not include individuals over 50 years of age, who are more likely to have decreased muscle mass. Fourth, we did not pair the US assessments with strength or performance-based outcome measures, which may reveal potential correlations with functional status, an area for future study. Finally, all examiners were experienced musculoskeletal sonographers, and results may differ for less experienced sonographers.

In conclusion, a robust normative reference for sonographic ratios of individual RTC muscle dimensions relative to adjacent muscles was established in an active, healthy, adult population of U.S. ADSMs, retirees, and dependents. While biological males have larger RTC muscle dimensions than females, their corresponding ratios are similar. The reliability of these ratios and potential influence of other demographic factors (e.g., participant height, body mass index, occupational specialty) on RTC muscle dimensions and corresponding ratios remain unknown and warrant further study. Additional research is needed to determine the clinical utility and predictive value of RTC muscle ratios for diagnosis of pathology (e.g., RTC tendon tear, denervation), compared to absolute RTC muscle dimensions alone.

Military-, service- and occupational specialty-specific demands should also be considered when assessing injury risk. For example, shoulder injuries are particularly prominent within the Special Operations Forces, who are exposed to higher loads during physical fitness and tactile training and deployment [18,19]. Future clinical studies should also explore how these ratios relate to functional performance, and how their application may influence return to duty decisions critical to warfighter readiness and maintaining a healthy, deployable force. [18]

## Data Availability

All data produced in the present study are available upon reasonable request to the authors.

## Acknowledgements

We thank Nelson Hager, MD, MS and John Persinger, BA, RDMS, RMSKS, CTT+ for their contributions to study protocol design and approval.

